# Spatial Accessibility to Hospital Capacity and the Concentration of Health-System Contributions in New Jersey

**DOI:** 10.64898/2026.09.02.26362067

**Authors:** Yuhao Zheng

## Abstract

**Background:** Hospital consolidation increasingly places multiple hospitals under common system control. However, spatial accessibility estimates alone do not indicate whether that accessibility is distributed across many health systems or concentrated within a few.

**Methods:** We used the enhanced two-step floating catchment area (E2SFCA) method to estimate tract-level spatial accessibility to staffed beds at general hospitals reachable within 30 minutes by car. For tracts with positive E2SFCA scores, we aggregated hospital contributions by health system and calculated a Herfindahl-Hirschman Index (HHI) from each system’s share of the tract’s total E2SFCA score. We used Spearman rank correlations to examine associations with tract-level sociodemographic characteristics.

**Results:** Of 163 reachable hospitals, 94 were outside New Jersey and accounted for 62.7% of staffed beds among these hospitals. Across 2,172 tracts, mean and median E2SFCA scores were 1.98 and 1.99 staffed hospital beds per 1,000 population, respectively. Among 2,161 tracts with positive E2SFCA scores, mean and median HHI values were 0.36 and 0.31. Using 2.0 for E2SFCA and 0.18 for HHI as descriptive reference values, 934 tracts had E2SFCA < 2.0 and HHI ≥ 0.18, while 655 had E2SFCA ≥ 2.0 and HHI ≥ 0.18. For seven of eight sociodemographic characteristics, correlations with E2SFCA and HHI were in opposite directions.

**Conclusions:** Considering E2SFCA and HHI together provides a community-level view of spatial accessibility and how that accessibility is distributed across health systems. This framework may inform assessments of hospital consolidation and proposed service or capacity changes.

## 1. Introduction

Access to health care varies by race and ethnicity, income, and neighborhood context (<u>Caraballo et al., 2022</u>; <u>Guo et al., 2022</u>; <u>Kirby & Yabroff, 2020</u>; <u>Tsui et al., 2020</u>; <u>Wolfe et al., 2020</u>). Part of this variation is spatial: provider capacity is not evenly distributed relative to population demand, and transportation constraints may make services difficult to reach. Spatial accessibility measures estimate potential geographic access by accounting for provider locations and capacity, population demand, and travel between communities and providers (<u>Guagliardo, 2004</u>). One widely used approach is the two-step floating catchment area (2SFCA) method (<u>Luo & Wang, 2003</u>), which estimates accessibility by relating provider capacity to population demand within overlapping catchments. The method and its extensions have been applied to examine geographic disparities in potential access to health care (<u>Campbell et al., 2025</u>; <u>Guagliardo, 2004</u>; <u>Kim et al., 2021</u>; <u>Luo, 2004</u>; <u>Mao & Nekorchuk, 2013</u>).

Spatial accessibility to hospital care is an important public health concern because general hospitals provide emergency care and time-sensitive inpatient services to surrounding communities. Prior studies have examined spatial accessibility to general hospitals and hospital-based services in a range of geographic settings in the United States (<u>Delamater, 2013</u>; <u>Delamater et al., 2019</u>; <u>Kang et al., 2020</u>; <u>Kim et al., 2021</u>; <u>McCrum et al., 2021</u>, <u>2022</u>; <u>Park et al., 2023</u>). However, less attention has been given to how different health systems contribute to a community’s spatial accessibility to hospital care. The share of U.S. community hospitals that were part of a health system rose from 58% in 2010 to 69% in 2023 (<u>Levinson et al., 2025</u>).

Hospital consolidation can place previously separate facilities under common ownership or control, allowing health systems to reorganize local services and capacity (<u>Bazzoli et al., 2002</u>; <u>Bogue et al., 1995</u>). When much of the hospital capacity available to a community is controlled by only a few health systems, decisions by those systems may have broader implications for local access to care.

This issue is particularly relevant in New Jersey. In 2025, 96% of the state’s general hospitals were part of health systems, and approximately half of the state’s staffed hospital beds were controlled by the three largest systems: RWJBarnabas Health, Hackensack Meridian Health, and Atlantic Health System (<u>DeLia et al., 2026</u>). Because New Jersey is embedded within the New York and Philadelphia metropolitan regions, hospitals in neighboring states may contribute substantially to the spatial accessibility of New Jersey communities.

This study examines two related dimensions of potential hospital access in New Jersey: tract-level spatial accessibility to staffed hospital beds and how that accessibility is distributed across health systems. We used the enhanced two-step floating catchment area (E2SFCA) method to estimate spatial accessibility, including contributions from reachable hospitals in surrounding states. For each tract with a positive E2SFCA score, we grouped hospital-level contributions by health system and calculated the Herfindahl–Hirschman Index (HHI) from each system’s share of the tract’s total E2SFCA score. Because this HHI is based on health-system contributions to tract-level spatial accessibility rather than conventional market shares within a defined geographic market, it should not be interpreted as a measure of market concentration or competition.

The study addresses three research questions. First, how does spatial accessibility to staffed hospital beds vary across New Jersey tracts? Second, among tracts with positive E2SFCA scores, how concentrated are hospital contributions to spatial accessibility across health systems? Third, how are E2SFCA scores and HHI values associated with tract-level sociodemographic characteristics?

## 2. Background

### 2.1 Measuring Spatial Accessibility to Health Care

Spatial accessibility describes potential access to health care based on the locations of providers and populations and the travel required to reach services. It represents the geographic dimension of access and is commonly distinguished from nonspatial dimensions, although geographic and nongeographic barriers often interact (<u>Joseph & Phillips, 1984</u>; <u>Khan, 1992</u>; <u>Luo & Qi, 2009</u>; <u>Meade et al., 1988</u>).

Measures of spatial accessibility differ in how they account for provider capacity, population demand, and travel impedance. Conventional approaches include provider-to-population ratios, nearest-facility distances, and gravity-based models. Provider-to-population ratios summarize supply relative to demand within fixed administrative areas, but they do not capture cross-boundary access or variation within those areas (<u>Guagliardo, 2004</u>; <u>McLafferty, 2003</u>). Nearest-facility measures calculate the distance to the closest provider but generally do not account for competing demand, provider capacity, or other reachable care options (<u>Guagliardo, 2004</u>; <u>Yang et al., 2006</u>). Gravity methods weight service opportunities by travel impedance, with more distant opportunities contributing less to the resulting accessibility estimate. However, gravity-based estimates can be sensitive to the specification of the impedance function and its parameter values (<u>Guagliardo, 2004</u>; <u>Kwan, 1998</u>; <u>McGrail & Humphreys, 2009</u>; <u>Talen & Anselin, 1998</u>; <u>Vale & Pereira, 2017</u>).

Two-step floating catchment area methods account for provider capacity and competing population demand within overlapping catchments. The original 2SFCA method (<u>Luo & Wang, 2003</u>) first calculates a provider-to-population ratio for each provider catchment and then sums the ratios associated with providers reachable from each population location. In the original formulation, all provider–population pairs within the catchment threshold receive the same distance weight, regardless of travel time. The enhanced two-step floating catchment area (E2SFCA) method instead applies distance-decay weights so that a provider’s contribution decreases as travel time increases (<u>Luo & Qi, 2009</u>). Later extensions have modified catchment definitions, distance-decay specifications, demand measures, transportation modes, and assumptions about provider choice and capacity (<u>Bauer & Groneberg, 2016</u>; <u>Bell et al., 2013</u>; <u>Delamater, 2013</u>; <u>Luo & Whippo, 2012</u>; <u>Mao & Nekorchuk, 2013</u>; <u>Wan et al., 2012</u>).

### 2.2 Hospital Consolidation and Health-System Contributions to Spatial Accessibility

Hospital consolidation often occurs through mergers and acquisitions (M&A), bringing previously separate hospitals or health systems under common ownership or control (<u>Cuellar & Gertler, 2003</u>; <u>Dranove & Lindrooth, 2003</u>). Much of the literature on hospital consolidation has examined its effects on prices, competition, efficiency, and quality of care (<u>Cooper et al., 2019</u>; <u>Gaynor et al., 2015</u>; <u>Gaynor & Town, 2012</u>; <u>Giancotti et al., 2017</u>; <u>Mariani et al., 2022</u>; <u>Vogt et al., 2006</u>). Less attention has been given to how consolidation relates to communities’ spatial accessibility and how different health systems contribute to that accessibility. Consolidation can allow a health system to reorganize services and capacity across hospitals in its local network. <u>Bogue et al. (1995)</u> describe two broad post-merger strategies: eliminating direct acute-care competitors and expanding acute-care networks. Under the first, an acquired hospital may close or discontinue acute-care functions; under the second, acute-care functions are retained as the system expands its network. <u>Bazzoli et al. (2002)</u> describe restructuring that can occur without hospital closure, including reductions in service duplication, consolidation of departments and programs, and staffing reductions. Studies have documented reductions in obstetric, surgical, diagnostic, primary care, and pediatric inpatient services at some acquired hospitals and the cessation of inpatient operations or complete closure at others (<u>Henke et al., 2021</u>; <u>Joseph et al., 2023</u>; <u>O’Hanlon et al., 2019</u>; <u>Wood et al., 2024</u>).

When several hospitals reachable from the same community belong to one health system, system-level changes can affect more than one source of capacity within that community’s reach. An E2SFCA score aggregates contributions from reachable hospitals without indicating whether those hospitals belong to the same or different health systems. We therefore use an HHI based on each system’s share of the tract’s E2SFCA score to characterize the degree of concentration across health systems.

## 3. Data and Methods

### 3.1 Study Area

The core study area included all 2020 Census tracts in New Jersey with a positive population in the 2020–2024 American Community Survey (ACS) 5-year estimates. The hospital supply included all general hospitals in New Jersey, New York, Pennsylvania, Delaware, and Maryland that were reachable within 30 minutes of at least one New Jersey tract. To capture competing demand from populations surrounding these hospitals, we defined a regional demand area comprising positive-population tracts in Connecticut, Delaware, Maryland, New Jersey, New York, and Pennsylvania that were within 30 minutes of at least one included hospital. Connecticut was included in the demand area because some of its tracts fell within the catchments of hospitals reachable from New Jersey tracts, although no Connecticut hospital was within 30 minutes of a New Jersey tract.

### 3.2 Data

#### 3.2.1 General Hospitals

Eligible facilities were short-term general acute care hospitals; psychiatric, rehabilitation, long-term acute care, and specialty hospitals were excluded. New Jersey general hospitals were identified using the <u>New Jersey Department of Health Acute-Care Facilities database</u>. No single source provided a complete list of eligible hospitals in the surrounding states. We therefore assembled and cross-checked a list of out-of-state general hospitals using <u>American Hospital Directory (AHD)</u> data, state hospital directories, hospital association records, hospital websites, annual reports, and public announcements. These sources were also used to determine whether hospitals were under common health-system ownership or control as of October 2025 and to assign each hospital to a health system.

Hospital capacity was measured primarily using total staffed beds reported in the 2025 <u>AHD</u> data, including general medical/surgical and special-care beds. When AHD provided only a consolidated facility-level count or lacked a hospital-specific value, we used the corresponding hospital-specific count from the <u>Leapfrog Hospital Survey</u>.

#### 3.2.2 Driving-Time Estimation Between Tract Centroids and Hospitals

We calculated driving times from population-weighted census tract centroids to geocoded eligible hospitals using the R5 routing engine through the r5r package in R. We built the automobile network from <u>Geofabrik</u> OpenStreetMap PBF extracts dated January 1, 2026, for Connecticut, Delaware, Maryland, New Jersey, New York, and Pennsylvania.

As tract centroids do not necessarily fall on routable road segments, we snapped each centroid to the nearest routable location on the R5 automobile network before calculating driving times. Among New Jersey tract origins, 96.1% were snapped within 0.25 km, 3.7% between 0.25 and 1 km, and 0.2% between 1 and 2 km; no New Jersey origin exceeded 2 km.

Routing used automobile mode with R5’s default static OpenStreetMap-derived speeds. Although January 20, 2026, at 10:00 a.m. was specified as the nominal departure time, automobile travel times were calculated from static network speeds rather than time-dependent traffic conditions. Accordingly, the estimates did not vary by departure time or day of the week and did not account for time-varying congestion.

We used a 30-minute travel-time catchment, consistent with the travel-time component of New Jersey’s network adequacy standard for acute care hospitals, which requires access within 20 miles or 30 minutes’ driving time, whichever is less, for 90% of covered persons in a county or service area (<u>N.J. Admin. Code § 11:24A-4.10, 2026</u>). We excluded tract–hospital pairs exceeding the 30-minute threshold. Hospitals reachable within 30 minutes of at least one positive-population New Jersey tract formed the final hospital sample.

#### 3.2.3 Tract-Level Sociodemographic Data

Total population and tract-level sociodemographic characteristics were obtained from the 2020–2024 ACS 5-year estimates. We included median household income; the percentages of residents below the poverty level, without health insurance, with a disability, aged 65 years or older, and not classified as non-Hispanic White alone; and the percentages of households without vehicle access and classified as limited English-speaking.

### 3.3 Methods

#### 3.3.1 E2SFCA Specification

We estimated spatial accessibility to staffed hospital beds using the enhanced two-step floating catchment area (E2SFCA) method developed by <u>Luo and Qi (2009</u>). The method first calculates a capacity-to-demand ratio for each hospital and then, for each tract, sums the travel-time-weighted ratios of reachable hospitals. We used the slow-decay weights reported by <u>Luo and Qi (2009)</u>. Driving times were divided into three bands: 0–10 minutes (*w_1_* = 1.00), more than 10–20 minutes (*w_2_* = 1.68), and more than 20–30 minutes (*w_3_* = 1.22), where denotes the weight assigned to band *m*.The same bands and weights were applied in both steps.

In the first step, we calculated a hospital-specific capacity-to-demand ratio *R_j_* for each included hospital *j*. Potential demand was represented by the travel-time-weighted population of positive-population tracts in the regional demand area within 30 minutes of hospital *j*.

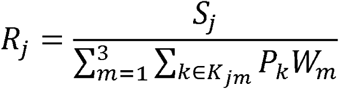

where *S_i_* is the staffed-bed capacity of hospital *j*, *p_k_* is the population of tract *k*, and *k_jm_* is the set of positive-population tracts *k* whose driving time to hospital *j* falls within travel-time band *m*.

In the second step, we calculated the unscaled E2SFCA score for each positive-population New Jersey tract by summing the travel-time-weighted values of hospitals reachable within 30 minutes:

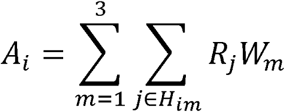

where *H_im_* is the set of hospitals whose driving time from tract *i* falls within travel-time band *m*. The term *R_j_W_m_* represents hospital *j*’s contribution to the spatial accessibility of tract *i*, with hospitals in more distant travel-time bands contributing less.

For reporting, we multiplied the unscaled E2SFCA scores by 1,000 and expressed the resulting scores as staffed hospital beds per 1,000 population.

#### 3.3.2 Health-System Contributions to Spatial Accessibility

To examine how health systems contributed to each tract’s spatial accessibility, we grouped the hospital-specific contributions to *A_i_* by system affiliation. Hospitals in the same health system were grouped together, while each independent hospital was treated as a separate single-hospital system.

For tract *i*, the contribution of health system to its unscaled E2SFCA score, *C_is_*, was defined as:

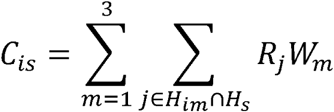

where *H_im_* is the set of hospitals whose driving time from tract *i* falls within travel-time band *m*, and *H_s_* is the set of hospitals belonging to health system *s*.

The health-system contributions sum to the tract’s total unscaled E2SFCA score:

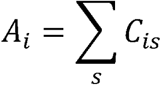

For tracts with *A_i_* > 0, the share of tract *i*’s spatial accessibility attributable to health system *s*, *p_is_*, was calculated as:

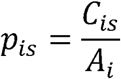

We then measured the concentration of these contributions using the Herfindahl– Hirschman Index (HHI):

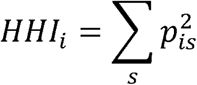

Higher HHI values indicate greater concentration of health-system contributions to a tract’s spatial accessibility.

#### 3.3.3 Descriptive Cross-Classification

We cross-classified tracts with positive E2SFCA scores using an E2SFCA reference value of 2.0 and an HHI reference value of 0.18, yielding four groups.

The E2SFCA reference value of 2.0 approximated the statewide mean (1.98) and median (1.99) and was used only as a descriptive reference near the center of the statewide distribution, not to define an adequate level of hospital capacity or spatial accessibility.

An HHI of 0.18 corresponds to 1,800 points on the conventional 0–10,000 scale. The 2023 Merger Guidelines classify markets with HHI values greater than 1,800 as highly concentrated (<u>U.S. Department of Justice & Federal Trade Commission, 2023</u>). For the descriptive cross-classification, we used 0.18 as a reference threshold and classified HHI values at or above 0.18 as the higher-concentration contribution group. In this study, HHI reflects the concentration of health-system contributions to a tract’s spatial accessibility, rather than firms’ market shares within a defined antitrust market.

### 3.4 Analysis of Sociodemographic Patterns

We used Spearman’s rank correlations to examine associations between each sociodemographic characteristic and the two tract-level measures: E2SFCA and HHI. Correlations involving E2SFCA were based on pairwise-complete observations among positive-population New Jersey tracts, whereas correlations involving HHI were additionally restricted to tracts with positive E2SFCA scores. Spearman’s rho is rank-based and does not require normally distributed variables (<u>Schober et al., 2018</u>). Because observations from nearby tracts may not be spatially independent, we treated the correlations as descriptive and did not report conventional significance tests. The correlations were not interpreted as causal relationships. Absolute coefficients below 0.10 were treated as negligible; values of 0.10–0.39 as weak, 0.40–0.69 as moderate, 0.70–0.89 as strong, and 0.90 or greater as very strong (<u>Schober et al., 2018</u>).

## 4. Results

### 4.1 Spatial Distribution of Reachable Hospitals and Their 30-Minute Catchments

The hospital sample comprised 163 general hospitals reachable within a 30-minute drive of at least one positive-population New Jersey tract. Of these hospitals, 69 were in New Jersey and 94 were outside the state: 58 in New York, 32 in Pennsylvania, 3 in Delaware, and 1 in Maryland. Five of the 163 hospitals were independent; the remaining 158 were part of health systems. Reachable hospitals and areas of higher kernel density of staffed hospital beds were concentrated around the New York City and Philadelphia metropolitan areas (Figures 1a and 1c). Out-of-state hospitals accounted for 62.7% of staffed beds in the hospital sample, compared with 37.3% for New Jersey hospitals.

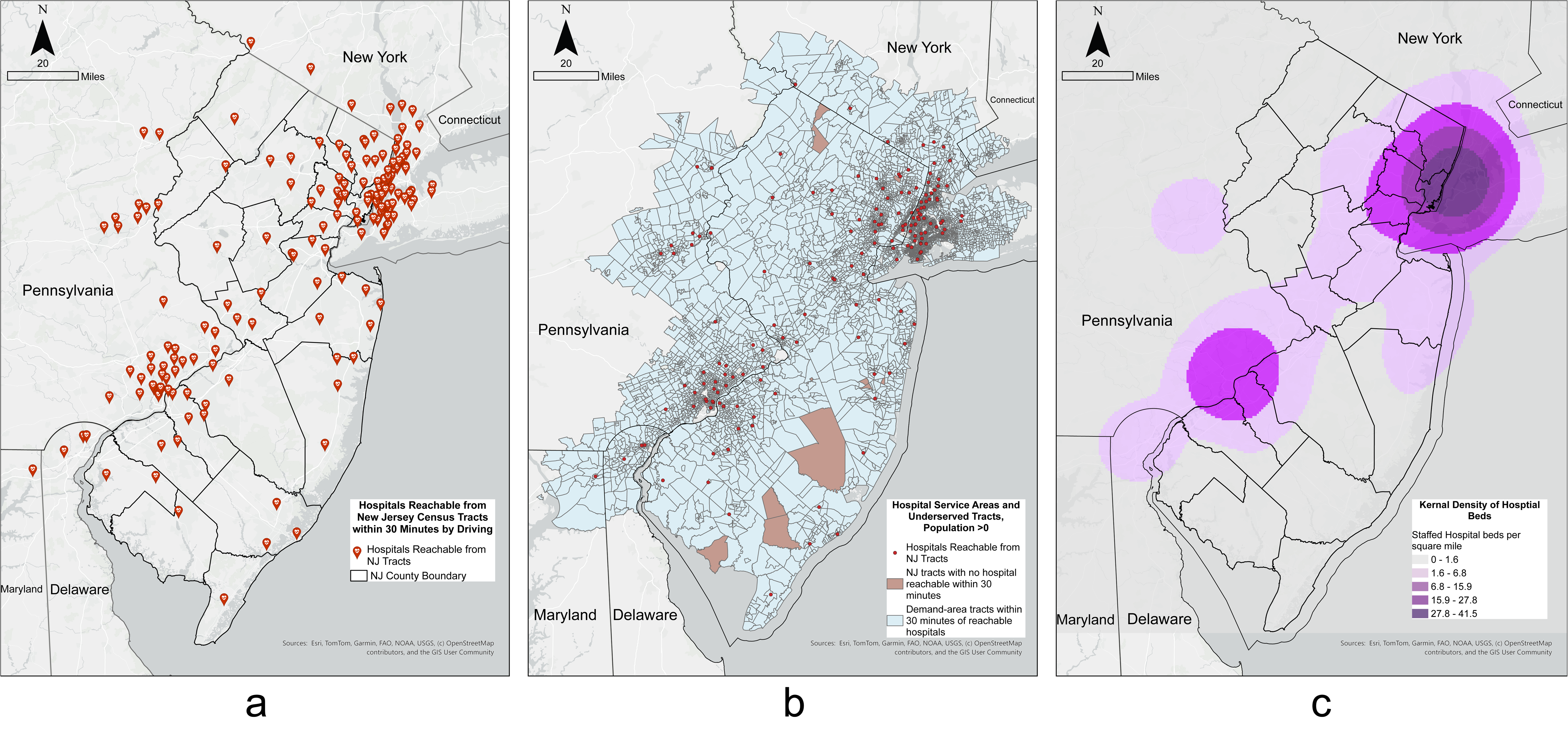

The regional demand area comprised 6,365 positive-population tracts within the combined 30-minute catchments of the 163 hospitals (Figure 1b). Of New Jersey’s 2,172 positive-population tracts, 2,161 were within a 30-minute drive of at least one eligible hospital. The remaining 11 tracts had a combined population of 46,867, representing 0.5% of the population across the study’s New Jersey tracts.

### 4.2 Spatial Accessibility and Concentration of Health-System Contributions

Across the 2,172 positive-population New Jersey tracts, the mean and median E2SFCA scores were 1.98 and 1.99, respectively, on a scale of staffed hospital beds per 1,000 population (Figure 2a). Scores at or above 2.0 occurred primarily in northeastern New Jersey and the Camden-Philadelphia area, with smaller clusters along the coast and in southern and northwestern New Jersey. Scores below 2.0 were widespread across the rest of the state.

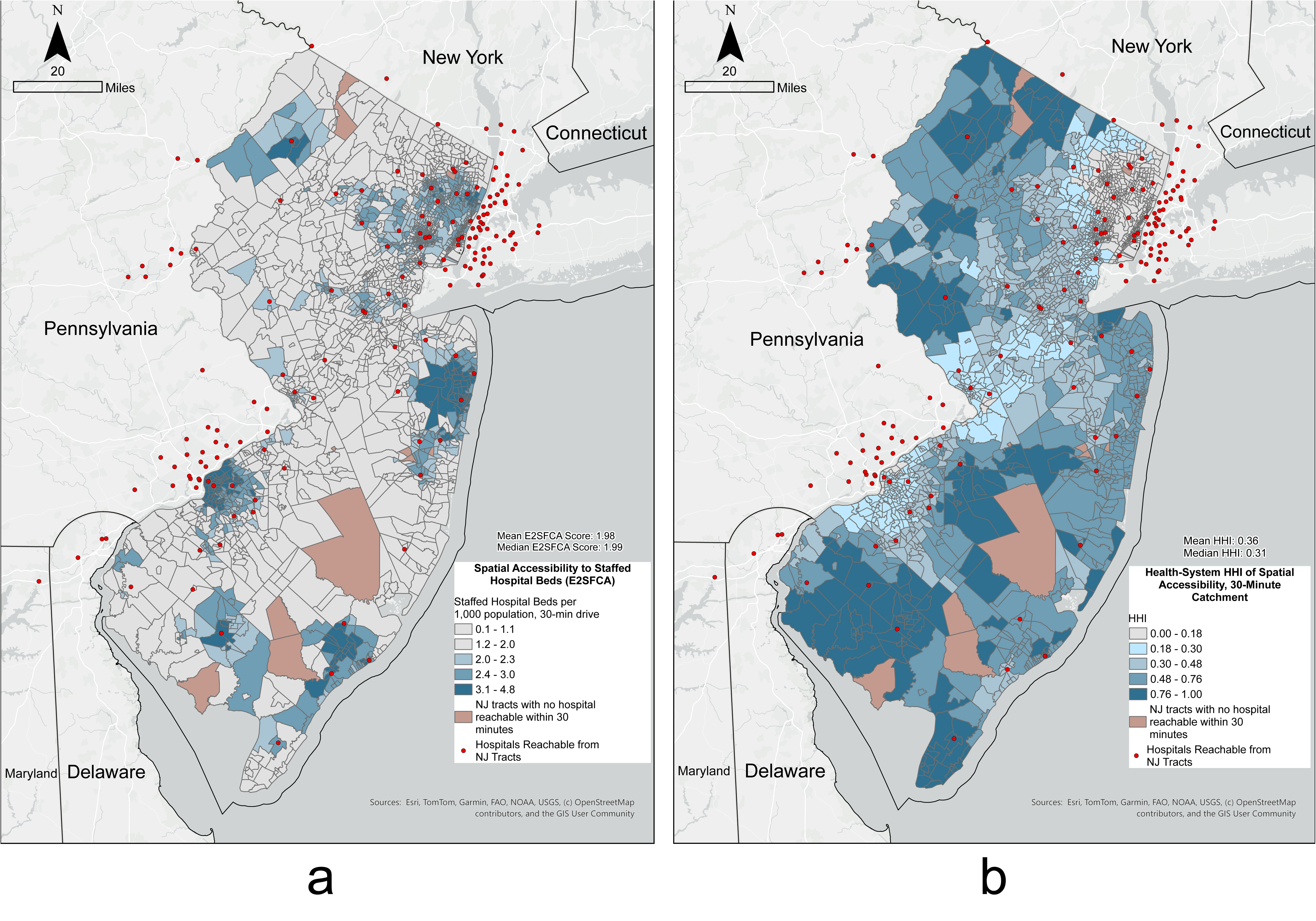

Among the 2,161 tracts with positive E2SFCA scores, the mean and median HHI values were 0.36 and 0.31, respectively (Figure 2b). HHI values below 0.18 occurred mainly in northeastern New Jersey and in smaller portions of the Camden-Philadelphia area and west-central New Jersey. Overall, 1,589 tracts (73.5%) had HHI values at or above 0.18.

Table 1 cross-classifies the 2,161 tracts using an E2SFCA reference value of 2.0 and an HHI reference value of 0.18. The largest category comprised 934 tracts (43.2%) with E2SFCA scores below 2.0 and HHI values at or above 0.18. An additional 655 tracts (30.3%) had E2SFCA scores at or above 2.0 and HHI values at or above 0.18. In total, 1,740 tracts (80.5%) fell below the E2SFCA reference value, met or exceeded the HHI threshold, or met both criteria.

**Table 1.**
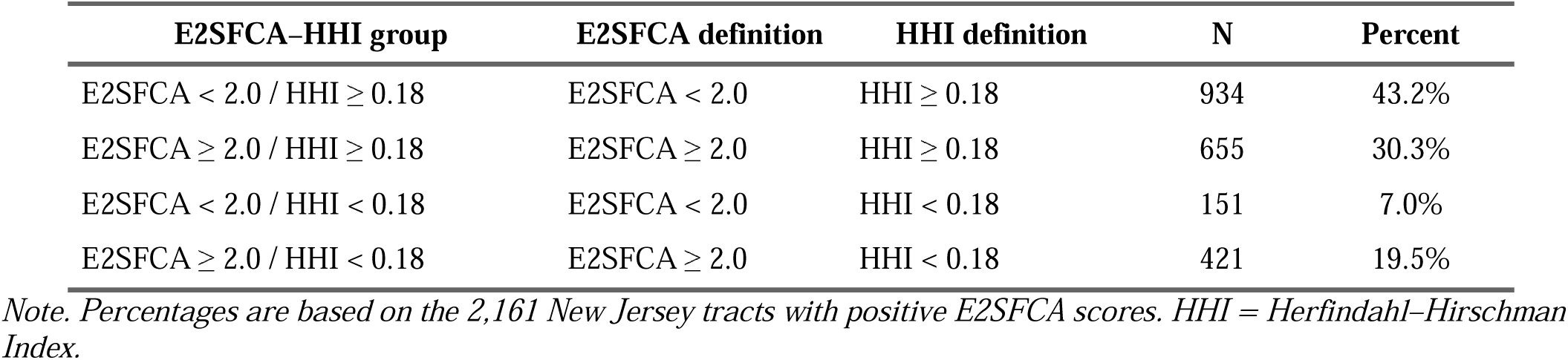
Distribution of New Jersey Tracts Across Four E2SFCA–HHI Groups.

| E2SFCA–HHI group | E2SFCA definition | HHI definition | N | Percent |
| --- | --- | --- | --- | --- |
| E2SFCA < 2.0 / HHI ≥ 0.18 | E2SFCA < 2.0 | HHI ≥ 0.18 | 934 | 43.2% |
| E2SFCA ≥ 2.0 / HHI ≥ 0.18 | E2SFCA ≥ 2.0 | HHI ≥ 0.18 | 655 | 30.3% |
| E2SFCA < 2.0 / HHI < 0.18 | E2SFCA < 2.0 | HHI < 0.18 | 151 | 7.0% |
| E2SFCA ≥ 2.0 / HHI < 0.18 | E2SFCA ≥ 2.0 | HHI < 0.18 | 421 | 19.5% |

*Note.* Percentages are based on the 2,161 New Jersey tracts with positive E2SFCA scores. HHI = Herfindahl–Hirschman Index.

### 4.3 Tract-Level Sociodemographic Correlates of E2SFCA and HHI

Table 2 presents tract-level Spearman rank correlations of eight sociodemographic characteristics with E2SFCA scores and HHI values. Pairwise-complete sample sizes ranged from 2,109 to 2,172 tracts for E2SFCA correlations and from 2,098 to 2,161 tracts for HHI correlations; HHI analyses were restricted to tracts with positive E2SFCA scores.

**Table 2.** Tract-Level Spearman Rank Correlations of Sociodemographic Characteristics with E2SFCA and HHI.

| Indicator | Spatial accessibility (E2SFCA):<br>Spearman $\rho$ (N) | Concentration of health-system<br>contributions (HHI): Spearman<br>$\rho$ (N) |
| --- | --- | --- |
| Median household income | -0.316 (N = 2,109) | 0.138 (N = 2,098) |
| Households without vehicle access | 0.365 (N = 2,166) | -0.411 (N = 2,155) |
| Population below poverty level | 0.313 (N = 2,168) | -0.225 (N = 2,157) |
| Population without health insurance | 0.301 (N = 2,168) | -0.287 (N = 2,157) |
| Limited English-speaking households | 0.271 (N = 2,166) | -0.468 (N = 2,155) |
| Population aged 65 years and older | -0.152 (N = 2,172) | 0.265 (N = 2,161) |
| Population with a disability | 0.074 (N = 2,168) | 0.135 (N = 2,157) |
| Residents not classified as non-Hispanic<br>White alone | 0.309 (N = 2,172) | -0.485 (N = 2,161) |

**Note.** Entries report Spearman’s $\rho$ , with the pairwise-complete N for each correlation shown in parentheses. HHI is undefined for tracts with zero E2SFCA scores. No significance stars or p-values are reported because the correlations are interpreted descriptively and observations from nearby tracts may not be spatially independent.

Seven of the eight E2SFCA correlations were weak in magnitude (|ρ| = 0.152–0.365), while the correlation with disability prevalence was negligible (ρ = 0.074). E2SFCA was positively correlated with poverty, lack of health insurance, the percentage of residents not classified as non-Hispanic White alone, lack of household vehicle access, and the percentage of limited-English-speaking households. It was negatively correlated with median household income and the percentage of residents aged 65 years or older.

For seven of the eight characteristics, the HHI correlation had the opposite sign from the corresponding E2SFCA correlation. The percentage of residents not classified as non-Hispanic White alone, the percentage of limited-English-speaking households, and the percentage of households without vehicle access had weak positive correlations with E2SFCA but moderate negative correlations with HHI (ρ = −0.485, −0.468, and −0.411, respectively). The remaining five HHI correlations were weak in magnitude (|ρ| = 0.135–0.287).

## 5. Discussion

Communities with similar E2SFCA scores can differ substantially in how hospital contributions to spatial accessibility are distributed across health systems. Where hospitals under the same system account for a large share of a community’s spatial accessibility, multiple reachable hospitals may still represent only a small number of health systems. E2SFCA and HHI therefore describe two different features of potential access: the level of spatial accessibility and how that accessibility is distributed across health systems.

For seven of the eight sociodemographic characteristics, correlations with E2SFCA and HHI had opposite signs. Several indicators of social disadvantage were associated with greater spatial accessibility and a less concentrated distribution of health-system contributions. This pattern may partly reflect the location of disadvantaged communities in metropolitan areas with greater hospital capacity and multiple health systems contributing to spatial accessibility. <u>Xie and Smart (2025)</u> likewise found greater spatial accessibility to primary care physicians in more socially disadvantaged areas of New Jersey. These findings, however, do not establish that residents of disadvantaged communities have better access to care overall. Geographic accessibility and availability represent only part of access to care <u>(Penchansky & Thomas, 1981)</u>. Barriers involving cost, insurance coverage, and scheduling may persist even where modeled accessibility is high and health-system contributions are relatively dispersed.

The substantial contribution of out-of-state hospitals shows that potential hospital access in New Jersey extends beyond state boundaries. Restricting the analysis to in-state hospitals would omit both capacity available across state lines and the competing demand from populations in neighboring states. For spatial accessibility measures, travel-based catchments therefore provide a more appropriate basis for defining spatial access than state boundaries alone.

In state planning and regulatory reviews of proposed hospital consolidation, ownership changes, closures, or capacity changes, E2SFCA and HHI could provide a community-level perspective on potential effects. A change in ownership could alter HHI even if hospital locations and capacity remained unchanged, whereas a hospital closure or change in capacity could affect both measures. Recalculating the measures under a proposed change could identify communities where spatial accessibility would decline, the concentration of health-system contributions would increase, or both. Such assessments would describe modeled changes under specified assumptions rather than causal effects or future utilization.

## 6. Limitations

Several limitations should be considered. First, we used total staffed hospital beds as a measure of hospital capacity. Staffed-bed capacity does not necessarily represent the number of beds actually available at a given time, particularly because occupancy can vary. We also did not distinguish among specific bed types or clinical services, which are not interchangeable. Future work could extend the framework to service-specific capacity where consistent data are available.

Second, the E2SFCA specification used here simplifies travel and hospital choice by applying a common 30-minute catchment and distance-decay function across hospitals and communities. Travel patterns and hospital-choice behavior may differ between emergency and elective care. The appropriate distance-decay specification may vary across services and settings (<u>Luo & Qi, 2009</u>). Our travel model also did not account for transportation availability, ambulance transport, time-varying congestion, weather conditions, referral patterns, hospital diversion status, or other logistical constraints that may affect access. E2SFCA measures potential spatial accessibility rather than realized access or utilization. Future work could use service-specific travel parameters or observed patient-flow data to refine these estimates.

Finally, the analysis is cross-sectional. The tract-level correlations are unadjusted and should not be interpreted as causal or individual-level relationships. We also did not account explicitly for spatial dependence among neighboring tracts. Future work could examine how both measures change over time as hospital organization and capacity change.

## 7. Conclusions

Spatial accessibility and the concentration of health-system contributions to that accessibility capture different features of potential hospital access. In New Jersey, communities with similar levels of spatial accessibility often differed substantially in how that accessibility was distributed across health systems, and sociodemographic characteristics showed different associations with the two measures. The substantial contribution of hospitals outside New Jersey also underscores the importance of defining hospital access using travel-based catchments rather than administrative boundaries. At the community level, E2SFCA and HHI can be used to examine how changes in hospital ownership, hospital capacity, or service capacity alter these dimensions of potential access to hospital care.

## Data Availability

All data produced in the present study are available upon reasonable request to the authors

## Declaration of Generative AI and AI-Assisted Technologies

During the preparation of this manuscript, the authors used ChatGPT (OpenAI) to assist with coding, literature searches, and language editing. The authors reviewed and verified all code, sources, citations, analyses, and text produced with its assistance and take full responsibility for the content of the manuscript.

## References

Bauer, J., & Groneberg, D. A. (2016). Measuring spatial accessibility of health care providers – Introduction of a variable distance decay function within the floating catchment area (FCA) method. PloS One, 11(7), Article e0159148. 10.1371/journal.pone.0159148

Bazzoli, G. J., LoSasso, A., Arnould, R., & Shalowitz, M. (2002). Hospital reorganization and restructuring achieved through merger. Health Care Management Review, 27(1), 7–20. 10.1097/00004010-200201000-00002

Bell, S., Wilson, K., Bissonnette, L., & Shah, T. (2013). Access to primary health care: Does neighborhood of residence matter? Annals of the Association of American Geographers, 103(1), 85–105. 10.1080/00045608.2012.685050

Bogue, R. J., Shortell, S. M., Sohn, M.-W., Manheim, L. M., Bazzoli, G., & Chan, C. (1995). Hospital reorganization after merger. Medical Care, 33(7), 676–686. 10.1097/00005650-199507000-00004

Campbell, J. E., Sambo, A. B., St. James, N., Willingham, K., Warren, H., Penaflor, J., & Doescher, M. P. (2025). Spatial accessibility for deploying mobile lung cancer screenings in a rural state using two-step floating catchment spatial analysis in Oklahoma, United States 2025. Preventive Medicine Reports, 60, Article 103290. 10.1016/j.pmedr.2025.103290

Caraballo, C., Ndumele, C. D., Roy, B., Lu, Y., Riley, C., Herrin, J., & Krumholz, H. M. (2022). Trends in racial and ethnic disparities in barriers to timely medical care among adults in the US, 1999 to 2018. JAMA Health Forum, 3(10), Article e223856. 10.1001/jamahealthforum.2022.3856

Cooper, Z., Craig, S. V., Gaynor, M., & Van Reenen, J. (2019). The price ain’t right? Hospital prices and health spending on the privately insured. The Quarterly Journal of Economics, 134(1), 51–107. 10.1093/qje/qjy020

Cuellar, A. E., & Gertler, P. J. (2003). Trends in hospital consolidation: The formation of local systems. Health Affairs, 22(6), 77–87. 10.1377/hlthaff.22.6.77

Delamater, P. L. (2013). Spatial accessibility in suboptimally configured health care systems: A modified two-step floating catchment area (M2SFCA) metric. Health & Place, 24, 30–43. 10.1016/j.healthplace.2013.07.012

Delamater, P. L., Shortridge, A. M., & Kilcoyne, R. C. (2019). Using floating catchment area (FCA) metrics to predict health care utilization patterns. BMC Health Services Research, 19(1), Article 144. 10.1186/s12913-019-3969-5

DeLia, D., Cantor, J., Wu, B., & Zheng, Y. (2026, July 13). Making sense of New Jersey hospital consolidation in a challenging policy environment. New Jersey State Policy Lab. https://policylab.rutgers.edu/publication/making-sense-of-new-jersey-hospital-consolidation-in-a-challenging-policy-environment/

Dranove, D., & Lindrooth, R. (2003). Hospital consolidation and costs: Another look at the evidence. Journal of Health Economics, 22(6), 983–997. 10.1016/j.jhealeco.2003.05.001

Gaynor, M., Ho, K., & Town, R. J. (2015). The industrial organization of health-care markets. Journal of Economic Literature, 53(2), 235–284. 10.1257/jel.53.2.235

Gaynor, M., & Town, R. (2012). The impact of hospital consolidation—Update (Policy Brief No. 9). The Synthesis Project, Robert Wood Johnson Foundation. https://www.healthwatchusa.org/HWUSA-References/editorial/20120600-rwjf73261.pdf

Giancotti, M., Guglielmo, A., & Mauro, M. (2017). Efficiency and optimal size of hospitals: Results of a systematic search. PloS One, 12(3), Article e0174533. 10.1371/journal.pone.0174533

Guagliardo, M. F. (2004). Spatial accessibility of primary care: Concepts, methods and challenges. International Journal of Health Geographics, 3(1), Article 3. 10.1186/1476-072X-3-3

Guo, J., Hernandez, I., Dickson, S., Tang, S., Essien, U. R., Mair, C., & Berenbrok, L. A. (2022). Income disparities in driving distance to health care infrastructure in the United States: A geographic information systems analysis. BMC Research Notes, 15(1), Article 225. 10.1186/s13104-022-06117-w

Henke, R. M., Fingar, K. R., Jiang, H. J., Liang, L., & Gibson, T. B. (2021). Access to obstetric, behavioral health, and surgical inpatient services after hospital mergers in rural areas: Study examines access to care in rural areas where hospitals have merged. Health Affairs, 40(10), 1627–1636. 10.1377/hlthaff.2021.00160

Joseph, A. M., Davis, B. S., & Kahn, J. M. (2023). Association between hospital consolidation and loss of pediatric inpatient services. JAMA Pediatrics, 177(8), 859–860. 10.1001/jamapediatrics.2023.1747

Joseph, A. E., & Phillips, D. R. (1984). Accessibility and utilization: Geographical perspectives on health care delivery. Harper & Row.

Kang, J.-Y., Michels, A., Lyu, F., Wang, S., Agbodo, N., Freeman, V. L., & Wang, S. (2020). Rapidly measuring spatial accessibility of COVID-19 healthcare resources: A case study of Illinois, USA. International Journal of Health Geographics, 19(1), Article 36. 10.1186/s12942-020-00229-x

Khan, A. A. (1992). An integrated approach to measuring potential spatial access to health care services. Socio-Economic Planning Sciences, 26(4), 275–287. 10.1016/0038-0121(92)90004-O

Kim, K., Ghorbanzadeh, M., Horner, M. W., & Ozguven, E. E. (2021). Identifying areas of potential critical healthcare shortages: A case study of spatial accessibility to ICU beds during the COVID-19 pandemic in Florida. Transport Policy, 110, 478–486. 10.1016/j.tranpol.2021.07.004

Kirby, J. B., & Yabroff, K. R. (2020). Rural–urban differences in access to primary care: Beyond the usual source of care provider. American Journal of Preventive Medicine, 58(1), 89–96. 10.1016/j.amepre.2019.08.026

Kwan, M. (1998). Space-time and integral measures of individual accessibility: A comparative analysis using a point-based framework. Geographical Analysis, 30(3), 191–216. 10.1111/j.1538-4632.1998.tb00396.x

Levinson, Z., Hulver, S., Godwin, J., & Neuman, T. (2025, February 19). Key facts about hospitals. KFF. https://www.kff.org/health-costs/key-facts-about-hospitals/

Luo, W. (2004). Using a GIS-based floating catchment method to assess areas with shortage of physicians. Health & Place, 10(1), 1–11. 10.1016/S1353-8292(02)00067-9

Luo, W., & Qi, Y. (2009). An enhanced two-step floating catchment area (E2SFCA) method for measuring spatial accessibility to primary care physicians. Health & Place, 15(4), 1100–1107. 10.1016/j.healthplace.2009.06.002

Luo, W., & Wang, F. (2003). Measures of spatial accessibility to health care in a GIS environment: Synthesis and a case study in the Chicago region. Environment and Planning B: Planning and Design, 30(6), 865–884. 10.1068/b29120

Luo, W., & Whippo, T. (2012). Variable catchment sizes for the two-step floating catchment area (2SFCA) method. Health & Place, 18(4), 789–795. 10.1016/j.healthplace.2012.04.002

Mao, L., & Nekorchuk, D. (2013). Measuring spatial accessibility to healthcare for populations with multiple transportation modes. Health & Place, 24, 115–122. 10.1016/j.healthplace.2013.08.008

Mariani, M., Sisti, L. G., Isonne, C., Nardi, A., Mete, R., Ricciardi, W., Villari, P., De Vito, C., & Damiani, G. (2022). Impact of hospital mergers: A systematic review focusing on healthcare quality measures. European Journal of Public Health, 32(2), 191–199. 10.1093/eurpub/ckac002

McCrum, M. L., Wan, N., Lizotte, S. L., Han, J., Varghese, T., & Nirula, R. (2021). Use of the spatial access ratio to measure geospatial access to emergency general surgery services in California. Journal of Trauma and Acute Care Surgery, 90(5), 853–860. 10.1097/TA.0000000000003087

McCrum, M. L., Wan, N., Han, J., Lizotte, S. L., & Horns, J. J. (2022). Disparities in spatial access to emergency surgical services in the US. JAMA Health Forum, 3(10), Article e223633. 10.1001/jamahealthforum.2022.3633

McGrail, M. R., & Humphreys, J. S. (2009). Measuring spatial accessibility to primary care in rural areas: Improving the effectiveness of the two-step floating catchment area method. Applied Geography (Sevenoaks), 29(4), 533–541. 10.1016/j.apgeog.2008.12.003

McLafferty, S. L. (2003). GIS and health care. Annual Review of Public Health, 24(1), 25–42. 10.1146/annurev.publhealth.24.012902.141012

Meade, M. S., Florin, J. W., & Gesler, W. M. (1988). Medical geography. Guilford Press. N.J.A.C. 11:24A-4.10 (2026). https://www.law.cornell.edu/regulations/new-jersey/N-J-A-C-11-24A-4-10

O’Hanlon, C. E., Kranz, A. M., DeYoreo, M., Mahmud, A., Damberg, C. L., & Timbie, J. (2019). Access, quality, and financial performance of rural hospitals following health system affiliation. Health Affairs, 38(12), 2095–2104. 10.1377/hlthaff.2019.00918

Park, J., Michels, A., Lyu, F., Han, S. Y., & Wang, S. (2023). Daily changes in spatial accessibility to ICU beds and their relationship with the case-fatality ratio of COVID-19 in the state of Texas, USA. Applied Geography, 154, Article 102929. 10.1016/j.apgeog.2023.102929

Penchansky, R., & Thomas, J. W. (1981). The concept of access: Definition and relationship to consumer satisfaction. Medical Care, 19(2), 127–140. 10.1097/00005650-198102000-00001

Schober, P., Boer, C., & Schwarte, L. A. (2018). Correlation coefficients: Appropriate use and interpretation. Anesthesia and Analgesia, 126(5), 1763–1768. 10.1213/ANE.0000000000002864

Talen, E., & Anselin, L. (1998). Assessing spatial equity: An evaluation of measures of accessibility to public playgrounds. Environment and Planning A, 30(4), 595–613. 10.1068/a300595

Tsui, J., Hirsch, J. A., Bayer, F. J., Quinn, J. W., Cahill, J., Siscovick, D., & Lovasi, G. S. (2020). Patterns in geographic access to health care facilities across neighborhoods in the United States based on data from the National Establishment Time-Series between 2000 and 2014. JAMA Network Open, 3(5), Article e205105. 10.1001/jamanetworkopen.2020.5105

U.S. Department of Justice, & Federal Trade Commission. (2023, December 18). *Merger guidelines*. https://search.ftc.gov/system/files/ftc_gov/pdf/P234000-NEW-MERGER-GUIDELINES.pdf

Vale, D. S., & Pereira, M. (2017). The influence of the impedance function on gravity-based pedestrian accessibility measures: A comparative analysis. Environment and Planning B: Urban Analytics and City Science, 44(4), 740–763. 10.1177/0265813516641685

Vogt, W. B., Town, R., & Williams, C. H. (2006). How has hospital consolidation affected the price and quality of hospital care? Robert Wood Johnson Foundation. https://search.issuelab.org/resources/6467/6467.pdf

Wan, N., Zou, B., & Sternberg, T. (2012). A three-step floating catchment area method for analyzing spatial access to health services. International Journal of Geographical Information Science, 26(6), 1073–1089. 10.1080/13658816.2011.624987

Wolfe, M. K., McDonald, N. C., & Holmes, G. M. (2020). Transportation barriers to health care in the United States: Findings from the National Health Interview Survey, 1997–2017. American Journal of Public Health, 110(6), 815–822. 10.2105/AJPH.2020.305579

Wood, C., Chen, X., Schpero, W., & Chatterjee, P. (2024). Acquisitions of safety-net hospitals from 2016–2021: A case series. Health Affairs Scholar, 2(6), Article qxae056. 10.1093/haschl/qxae056

Xie, Y., & Smart, M. (2025). Attractive accessibility: Exploring disparities in attributes of primary care physicians in New Jersey. Health & Place, 95, Article 103535. 10.1016/j.healthplace.2025.103535

Yang, D.-H., Goerge, R., & Mullner, R. (2006). Comparing GIS-Based methods of measuring spatial accessibility to health services. Journal of Medical Systems, 30(1), 23–32. 10.1007/s10916-006-7400-5

